# Non-pharmaceutical interventions and the emergence of pathogen variants

**DOI:** 10.1101/2021.05.27.21257938

**Authors:** Ben Ashby, Cameron A. Smith, Robin N. Thompson

## Abstract

Non-pharmaceutical interventions (NPIs), such as social distancing and contact tracing, are important public health measures that can reduce pathogen transmission. In addition to playing a crucial role in suppressing transmission, NPIs influence pathogen evolution by mediating mutation supply, restricting the availability of susceptible hosts, and altering the strength of selection for novel variants. Yet it is unclear how NPIs might affect the emergence of novel variants that are able to escape pre-existing immunity (partially or fully), are more transmissible, or cause greater mortality. We analyse a stochastic two-strain epidemiological model to determine how the strength and timing of NPIs affects the emergence of variants with similar or contrasting life-history characteristics to the wildtype. We show that, while stronger and timelier NPIs generally reduce the likelihood of variant emergence, it is possible for more transmissible variants with high cross immunity to have a greater probability of emerging at intermediate levels of NPIs. This is because intermediate levels of NPIs allow an epidemic of the wildtype that is neither too small (facilitating high mutation supply), nor too large (leaving a large pool of susceptible hosts), to prevent a novel variant becoming established in the host population. However, since one cannot predict the characteristics of a variant, the best strategy to prevent emergence is likely to be implementation of strong, timely NPIs.

## Introduction

Understanding the impact of interventions for infectious disease control on pathogen evolution is a major challenge at the interface of public health and evolutionary biology (Porco *et al*., 2005; Read *et al*., 2015; Day *et al*., 2020). To date, most theoretical explorations of how interventions mediate pathogen evolution have focused on the impact of vaccinations on antigenic evolution (McClean, 1995; Wilson *et al*., 1998; van Boven *et al*., 2005), changes in pathogen population structure (Lipsitch, 1997; Watkins *et al*., 2015), and virulence evolution (Gandon *et al*., 2001, 2003; André & Gandon, 2006; Ganusov & Antia, 2006; Gandon & Day, 2007; Williams & Day, 2008; Miller & Metcalf, 2019). For example, imperfect vaccines have been shown both theoretically (Gandon *et al*., 2001, 2003) and empirically (Read *et al*., 2015) to select for higher virulence by allowing vaccinated hosts to continue transmitting while infected (for a discussion of when this might occur see Bull & Antia, 2022). Certain interventions may therefore have unintended consequences for pathogen evolution and long-term negative impacts on the host population. Recently, there has been renewed interest in the effects of vaccination programmes on pathogen evolution and subsequent transmission during the COVID-19 pandemic (Bieniasz, 2021; Cobey *et al*., 2021; Gog *et al*., 2021; Saad-Roy *et al*., 2021; Sah *et al*., 2021; Thompson *et al*., 2021). This includes the potential for new pathogen variants (otherwise referred to as “mutants” or “strains” in the literature), which may be more transmissible, more virulent, or able to escape naturally- or vaccine-induced immunity. However, while the impact of vaccination on pathogen evolution has been the subject of sustained theoretical interest, there has been relatively little attention directed at the effects of non-pharmaceutical interventions on pathogen evolution (Hartfield & Alizon, 2014; Day *et al*., 2020; Gurevich *et al*., 2021).

NPIs, including social distancing, improved hygiene practices, school closures, lockdowns, quarantining exposed or isolating infected individuals, contact tracing, and various other measures, are important tools for managing outbreaks of infectious diseases. NPIs are particularly crucial during the early stages of an epidemic when pharmaceutical interventions (including vaccines) may not be available (e.g. for a novel pathogen). Although various NPIs have been used during previous epidemics of seasonal and pandemic influenza, Ebola, and Zika, among others, NPIs were almost universally adopted (to an extent not seen before) across the globe during the COVID-19 pandemic. Some NPIs, such as mask-wearing and national lockdowns, have been credited with drastically reducing cases and bringing epidemics under control, both prior to and in conjunction with vaccination programmes (Thompson *et al*., 2020; Li *et al*., 2021; Liu *et al*., 2021; Moore *et al*., 2021). Given their widespread adoption during the COVID-19 pandemic, it is likely that NPIs will feature heavily in public health responses to future epidemics or pandemics, and therefore we must improve our understanding of how NPIs affect both pathogen transmission and evolution.

The emergence of a novel pathogen variant occurs in two stages. First, the variant must be generated through mutation or recombination (“appearance”). Mutation supply governs the appearance of a new variant and depends on the rate at which the pathogen replicates in the host population (determined by the number of infected hosts), the mutation rate, and the number of mutations required to generate the variant. By reducing opportunities for transmission, NPIs effectively lower the rate at which the pathogen replicates, and hence lower the mutation supply. NPIs should therefore always make the appearance of novel variants less likely by restricting mutation supply.

If a variant does appear, then the second stage for emergence requires sustained transmission between hosts (“establishment”). It should be noted that many variants are likely to appear that have little or no impact on pathogen transmission or virulence. Variants may appear but remain undetected as they are unable to become established due to selection (if the variant is less fit than the wildtype) (Kucharski & Gog, 2012) or stochastic extinction (if by chance the variant dies out before it can infect many hosts) (Ferguson & Galvani, 2003; Abu-Raddad & Ferguson, 2004; Gog, 2008). NPIs are likely to have more complex effects on the establishment phase as they influence the strength of selection, the likelihood of stochastic extinction, and the availability of susceptible hosts. For example, stronger NPIs will slow the rate at which a variant can spread and will increase the likelihood of stochastic extinction, but will also change how immunity accumulates in the host population.

Previous theory has demonstrated the importance of the relationship between the appearance and establishment phases of variant emergence in the absence of NPIs (Lourenço & Recker, 2010; Hartfield & Alizon, 2014). In particular, Hartfield and Alizon (2014) analytically derived the probability of a strongly adapted variant with full cross-immunity (*R*_0_ ≫1) emerging from a weakly adapted strain (*R*_0_ ≈1), accounting for depletion of susceptible hosts, mutation supply, and stochastic extinction (*R*_0_ is the basic reproduction number, which represents the expected number of secondary infections caused by a pathogen in an otherwise susceptible population). The ongoing depletion of susceptible hosts was shown to be especially important for suppressing the emergence of a novel variant. Arinaminpathy & McLean (2009) modified a modelling approach by Antia *et al*. (2003) to investigate the emergence and establishment of a pathogen that is adapting to transmit more effectively in humans, providing a mathematical approach for monitoring outbreaks for signs of pathogen emergence, again in the absence of NPIs.

Here, we analyse a stochastic model of pathogen evolution to explore the effects of NPIs on the emergence of novel variants which may have no, partial, or full cross-immunity. We show how the strength and timing of NPIs, along with the life-history characteristics of the variant, affect its emergence and impact on the host population.

## Methods

To explore the key factors underlying the emergence of novel variants in a simple setting, we model infectious disease dynamics in a well-mixed, homogeneous host population over a relatively short period, ignoring host births and natural mortality for simplicity (see Fig. 1 for a model schematic). We restrict our model to two strains, a wildtype (*w*) and a variant (*ν*). The transmission rates of both strains are equally affected by NPIs, as determined by the parameter *r* (0 ≤ *r* ≤ 1). We assume that, for each strain, the baseline transmission rate is *β*_*i*_ (*i* ∈ {*w, ν*}), the disease-associated mortality rate (virulence) for single infections is *α*_*i*_, and the average infectious period is 1/*γ*.

**Figure 1.**
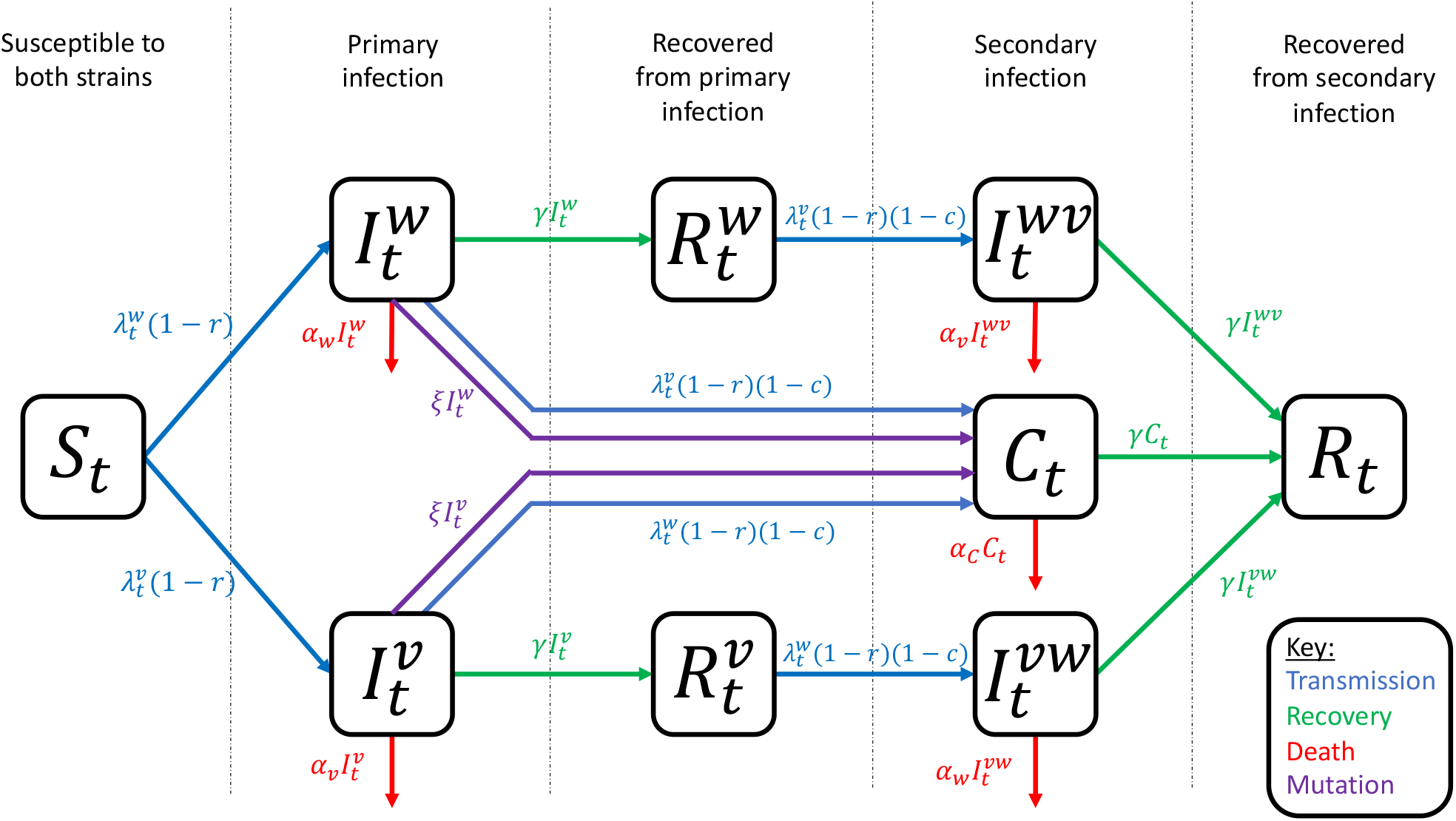
Model schematic. Arrows show transitions between classes at the given rates, with parameters as described in the main text.

Following a primary infection, an individual is assumed to be fully immune to the strain by which they were infected, and to have acquired partial cross-immunity, *c*, to the other strain (0 ≤ *c* ≤ 1). Thus, when *c* = 0 there is no cross-immunity between strains and when *c* = 1 there is full cross-immunity. Cross-immunity is assumed to only reduce a host’s susceptibility to infection by the other strain, such that the probability of successful infection at each potential infectious contact is reduced by a factor of (1 − *c*) (Thompson *et al*., 2019b). The order in which a host is infected does not affect the outcome of infection by the first or second strain (i.e. the virulence of a strain is the same regardless of whether the host experiences a primary or secondary infection). We assume that the variant differs from the wildtype at a single genetic locus, and that mutations occur between strains at overall rate *ξ* in each infected host, leading to coinfections by both strains. The virulence for coinfections is assumed to be the average of the wildtype and variant, 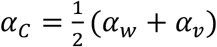.

At time, *t*, we use the following notation: *S*_*t*_ is the number of hosts that are fully susceptible to both the wildtype and the variant (initially all individuals are susceptible to both strains), 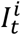 is the number of primary infections by strain *i* (i.e. individuals who are currently infected by strain *i* and have never previously been infected by either strain), 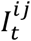 is the number of secondary infections by strain *j* following recovery from primary infection by strain *i, C*_*t*_ is the number of coinfections, 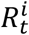 is the number of individuals who have recovered from a primary infection by strain *i* (and are immune to strain *i*, and not yet infected by the other strain), and *R*_*t*_ the number of individuals who have recovered from both strains (and are therefore immune to both strains). In the absence of NPIs, the per-capita force of infection on fully susceptible hosts for strain *i* is denoted 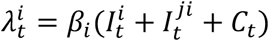 (*i* ≠ *j* throughout).

We assume that NPIs are either triggered (i) at the start of the epidemic and remain in place for the rest of a simulation, or (ii) when the total disease prevalence, 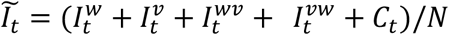, exceeds a threshold of *∈*_*on*_ > 0 and remain in place until 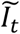 falls below a second threshold, *∈*_*off*_ > 0, where *∈*_*on*_ > *∈*_*off*_. Primary infections by strain *i* occur at per-capita rate 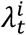 before NPIs are triggered and 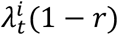 after NPIs are triggered. Similarly, secondary infections and coinfections (not arising from mutation) occur at per-capita rates 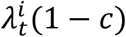 and 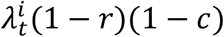 before and after NPIs are triggered, respectively. Assuming mutations are relatively rare (*ξ* ≪ *γ*), the effective (or time-varying) reproduction number (Cori *et al*., 2013; Thompson *et al*., 2019a) of strain *i* at time *t* in the absence of restrictions is given by:

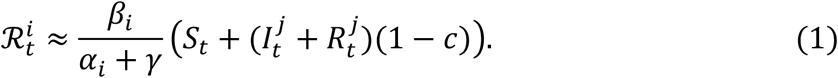

We run simulations of the model using the direct method version of the Gillespie stochastic simulation algorithm (Gillespie, 1977) using a population size of *N* = 100,000 and initial condition 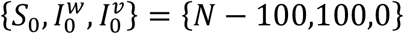, with 1,000 simulations per parameter set unless otherwise stated. Since here we are only interested in the emergence of a new variant, we fix the following parameters as they do not qualitatively change our results: 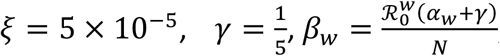, and 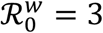. We choose *β*_*ν*_ such that the variant is less 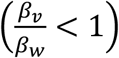 equally 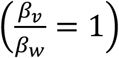, or more 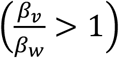 transmissible than the wildtype. We also vary whether NPIs are on for the duration of an epidemic or have trigger thresholds *ξ∈*_*on*_ = 0.01, *∈*_*off*,,_ = 0.0021, the level of cross-immunity between the wildtype and variant (0 ≤ *c* ≤ 1), the strength of NPIs (0 ≤ *r* ≤ 1), the virulence of the wildtype 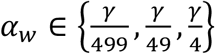 giving an infection fatality rate for the wildtype of 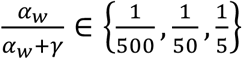, and the relative virulence of the variant 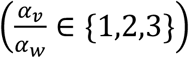. We define emergence by concluding that the variant has emerged if the frequency of infections caused by the variant exceeds 10% at any time during a simulation. Simulations are terminated when the number of hosts infected reaches 0.

## Results

### WHEN IS THE VARIANT FITTER THAN THE WILDTYPE?

We consider a scenario in which the variant is initially rare relative to the wildtype. The variant is fitter than the wildtype when 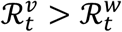, which requires:

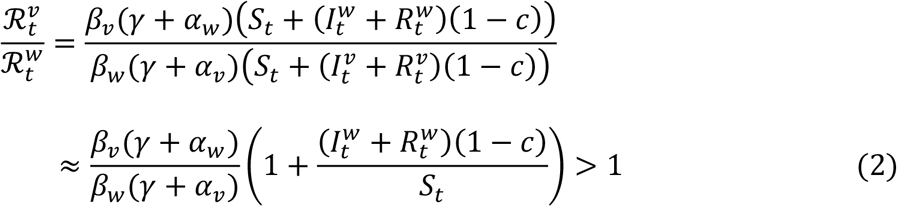

Hence, a variant is always fitter when it is at least as transmissible as the wildtype, has virulence than or equal to the wildtype, and cross immunity is less than complete (*c* < 1). However, a less transmissible variant may also be fitter provided the wildtype has already infected a sufficient proportion of the population and there is not full cross immunity.

Clearly, when *c* < 1 the fitness of the variant will increase relative to the wildtype as the pool of susceptible hosts for the latter is depleted. Note that NPIs do not directly affect whether a variant is fitter than the wildtype since there is no differential effect on transmission. NPIs will, however, affect mutation supply (and hence the appearance of the variant) and the rate at which the variant can spread (establishment) by modifying the transmission rate and the availability of susceptible hosts.

### TIMELY AND PERSISTENT NPIS

We initially consider the case when NPIs are triggered immediately and remain in place throughout the epidemic (Fig. 2). When NPIs are sufficiently strong to prevent the wildtype from initially spreading 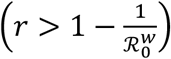, then unless the mutation rate is very high, it is unlikely that the variant will appear before the wildtype is driven extinct. Thus, the probability of a variant emerging is close to 0 whenever 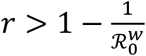 regardless of transmissibility or cross immunity (Fig. 2i-l). When *r* < min 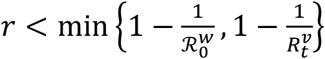, the probability of a variant emerging depends on the relative transmissibility of the variant, the strength of cross immunity, and the strength of NPIs. When cross immunity and NPIs are both relatively weak, there is a high probability of variants emerging (Fig. 2i-l). This is because there is a high mutation supply (weak restrictions) to generate the variant and a large pool of susceptible individuals to exploit.

**Figure 2.**
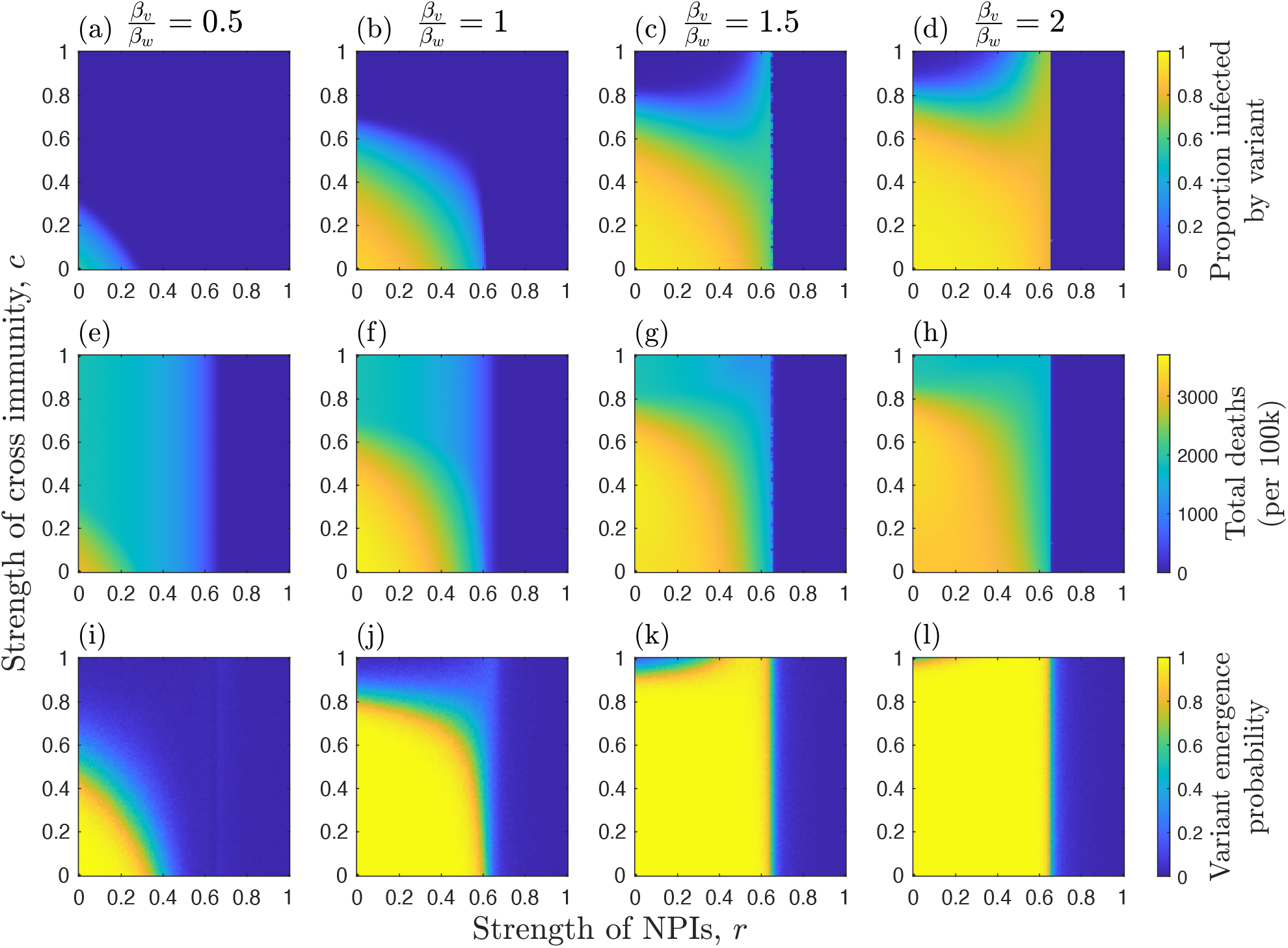
Effects of the strength of NPIs (*r*), strength of cross immunity (*c*), and relative transmissibility of the variant (*β*_*ν*_*β*_*w*_; as indicated at the top of each column) on: (a)-(d) median proportion of hosts infected by the variant; (e)-(h) median total deaths (per 100k) for both strains; and (i)-(l) the probability of the variant emerging (reaching a frequency of at least 0.1). Each measure is calculated over the full duration of each simulation. NPIs are triggered at the start of each simulation and remain in place throughout. Other parameters as described in the main text, with: 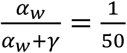 and *α*_*ν*_ = *α*_*w*_.

When the variant is less or equally as transmissible as the wildtype, there is an inverse effect of cross immunity and NPIs on the probability of the variant emerging: weaker NPIs require stronger cross immunity, and vice versa, to prevent the variant emerging (Fig. 2i-j). When the variant is sufficiently more transmissible, however, it has a high probability of emerging even when cross immunity is strong (Fig. 2k-l). Most notably, the variant is more likely to emerge for intermediate NPIs when cross-immunity is very high or complete (*c* ≈1). This is because, when NPIs are weak (*r* ≪ 1), there is a large outbreak of the wildtype, which leads to a high mutation supply but also rapidly depletes the pool of hosts for the variant due to cross-immunity (Fig. 3a, d). Thus, while a variant is likely to appear, it is unlikely to spread widely in the population due to herd immunity (Ashby & Best, 2021). When NPIs are stronger (but not too strong) the wildtype causes a smaller outbreak, leading to a lower build-up of cross immunity in the population while still maintaining a sufficient mutation supply for the variant to appear with high probability (Fig. 3b, e). This creates just the right conditions to allow the variant to sweep into the population. As a result of this phenomenon, it is possible for total deaths (for both strains) to peak at intermediate NPIs when the variant is both more transmissible and more deadly than the wildtype (Fig. 4). These results are robust to variation in virulence (Fig. S1-S3).

**Figure 3.**
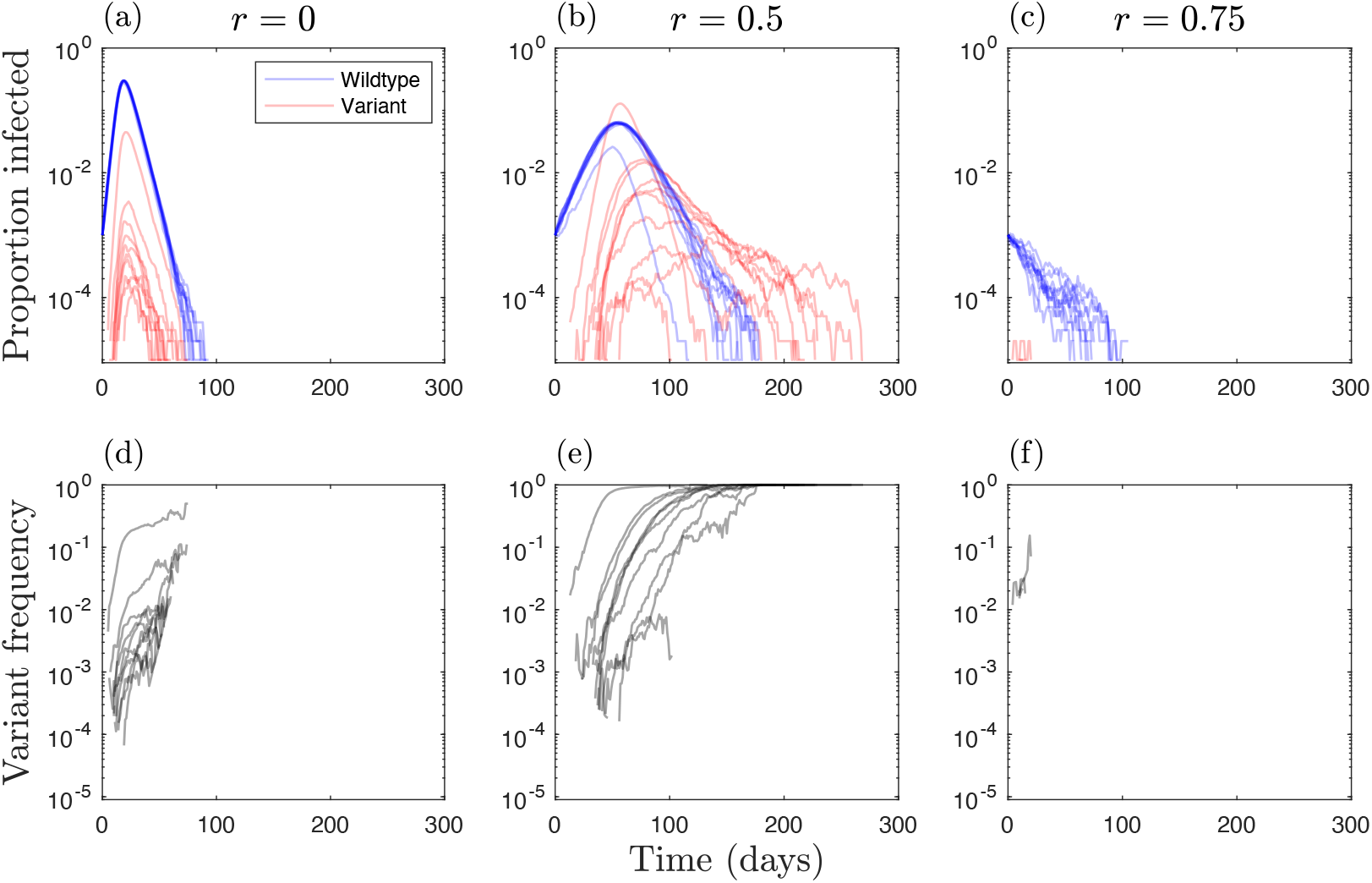
Simulations with full cross immunity and a 50% more transmissible variant 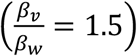 for different levels of NPIs (triggered at the start of the simulation and in place throughout): (a, d) no NPIs (*r* = 0); (b, e) intermediate NPIs (*r* = 0.5); (c, f) strong NPIs (*r* = 0.75). (a-c) Proportion of the entire population currently infected by the wildtype (blue) and by the variant (red), with coinfecteds contributing to both counts. (d-f) Frequency of the variant. Simulations were terminated when the pathogen went extinct. Other parameters as described in the main text, with: 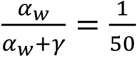 and *α*_*ν*_ = *α*_*w*_.

**Figure 4.**
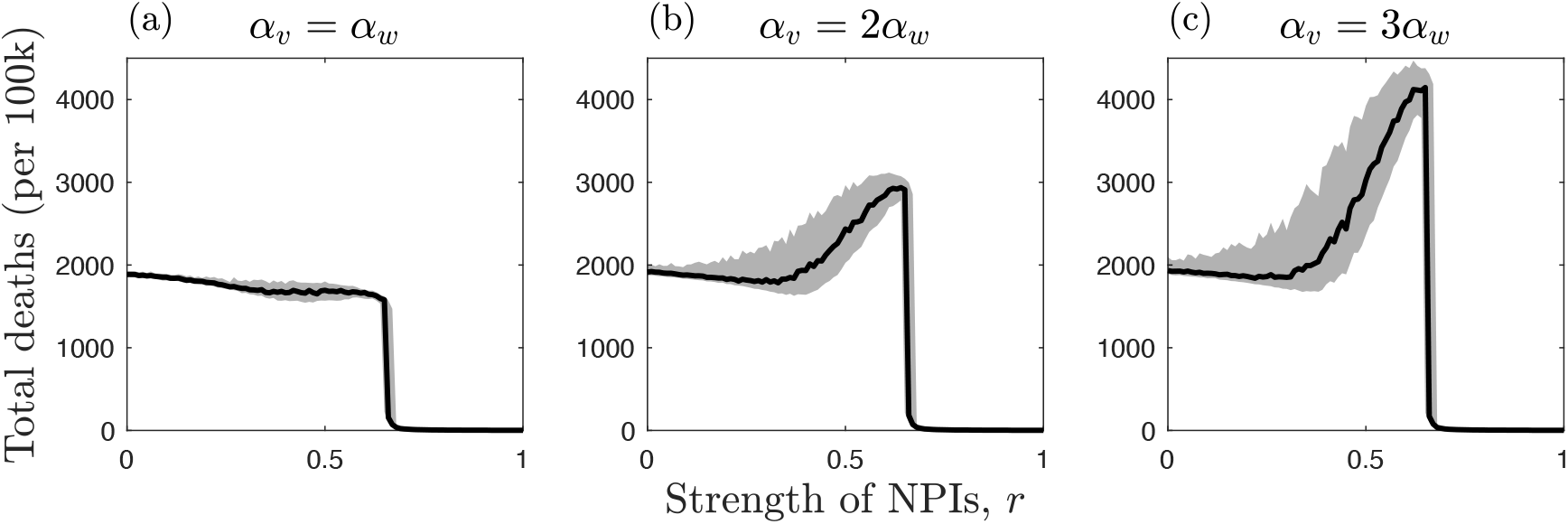
Effects of NPIs (*r*) and the relative virulence (*α*_*ν*_/*α*_*w*_) of a twice as transmissible variant (*β*_*ν*_/*β*_*w*_ = 2) on total deaths (per 100k) when there is full cross immunity (*c* = 1) and NPIs are introduced at the start of the simulation and remain in place throughout. Plots show the median number of deaths (black) along with the upper and lower quartiles (grey). Other parameters as described in the main text with 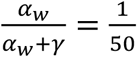.

### NPIS WITH TRIGGER THRESHOLDS

We now consider the case where NPIs are only triggered when disease prevalence is above a non-zero threshold (*∈*_*on*_ = 0.01) and are removed when disease prevalence falls below a second threshold (*∈*_*off*,,_ = 0.002) (Fig. 5). Hence, NPIs are triggered when 1% of the host population is infected by either strain, and are removed when only 0.2% are infected. There are three notable effects of having triggering thresholds for NPIs. First, since NPIs are not in place from the start, a non-negligible number of deaths now occur over the full range of NPI strengths (Fig. 5e-h). Second, as the initial mutation supply is no longer curtailed by NPIs, variants can emerge for 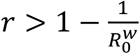 (Fig. 5j-l). Third, more transmissible variants which experience sufficiently high cross immunity with the wildtype (*c* ≈1) are still most likely to emerge at intermediate NPIs, but now the variant remains highly likely to emerge when NPIs are strong but fall below the relaxation threshold *∈*_*off*_ (compare Fig. 2k-l and Fig. 5k-l). However, if the variant is also more virulent, then deaths still peak when NPI intensity is intermediate (Fig. 6). These results are robust to variation in virulence (Fig. S4-S6).

**Figure 5.**
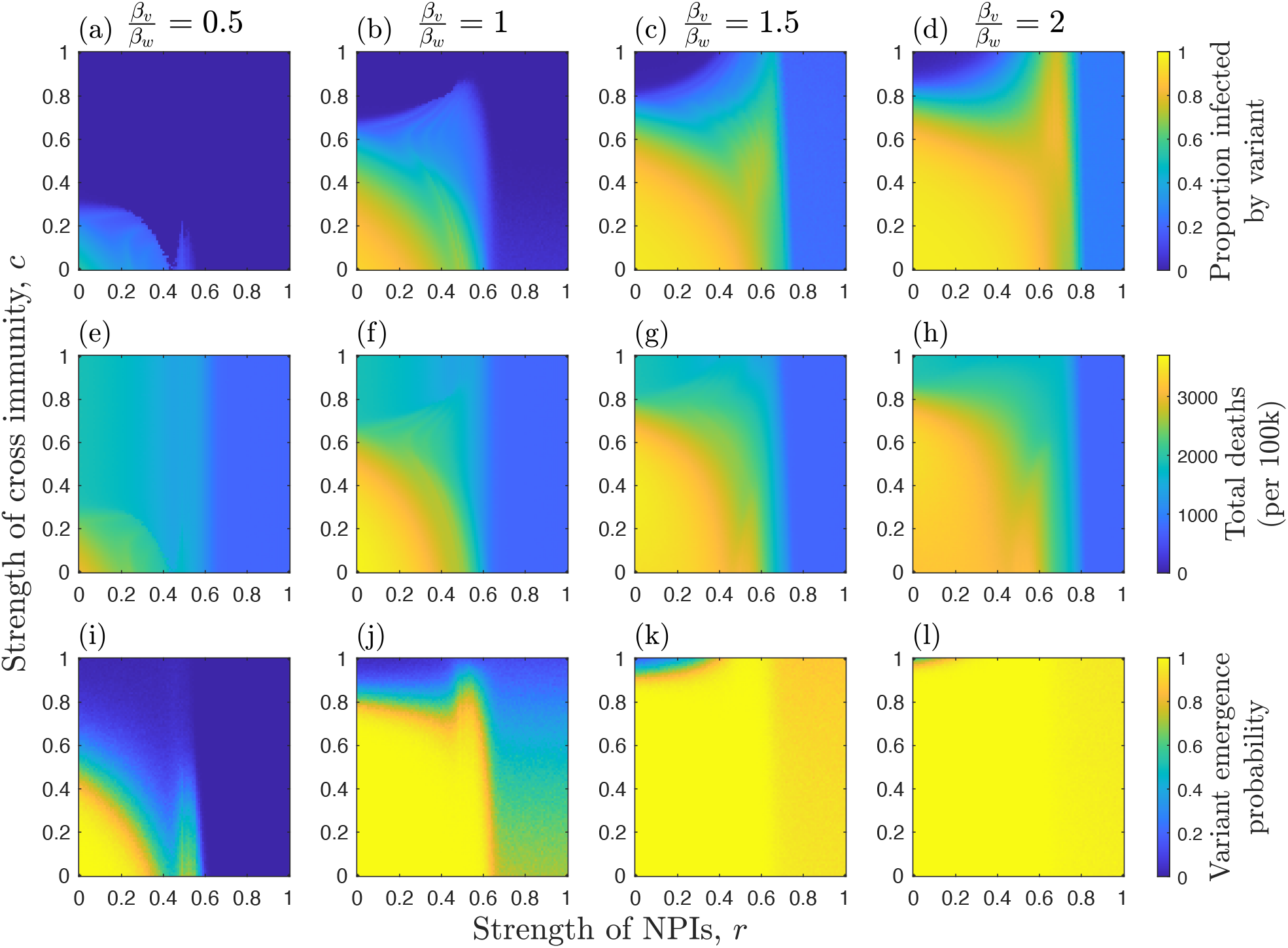
Effects of the strength of NPIs (*r*) when there are NPI trigger thresholds, strength of cross immunity (*c*), and relative transmissibility of the variant (*β*_*ν*_/*β*_*w*_; as indicated at the top of each column) on: (a)-(d) median proportion of hosts infected by the variant; (e)-(h) median total deaths (per 100k) for both strains; and (i)-(l) the probability of the variant emerging (reaching a frequency of at least 0.1). The NPI trigger thresholds are *∈*_*on*_ = 0.01 and *∈*_*off*,,_ = 0.002 (i.e. NPIs are triggered when 1% of the host population is infected by either strain, and are removed when only 0.2% are infected). Other parameters as described in the main text, with: 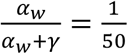and *α*_*ν*_ = *α*_*w*_.

**Figure 6.**
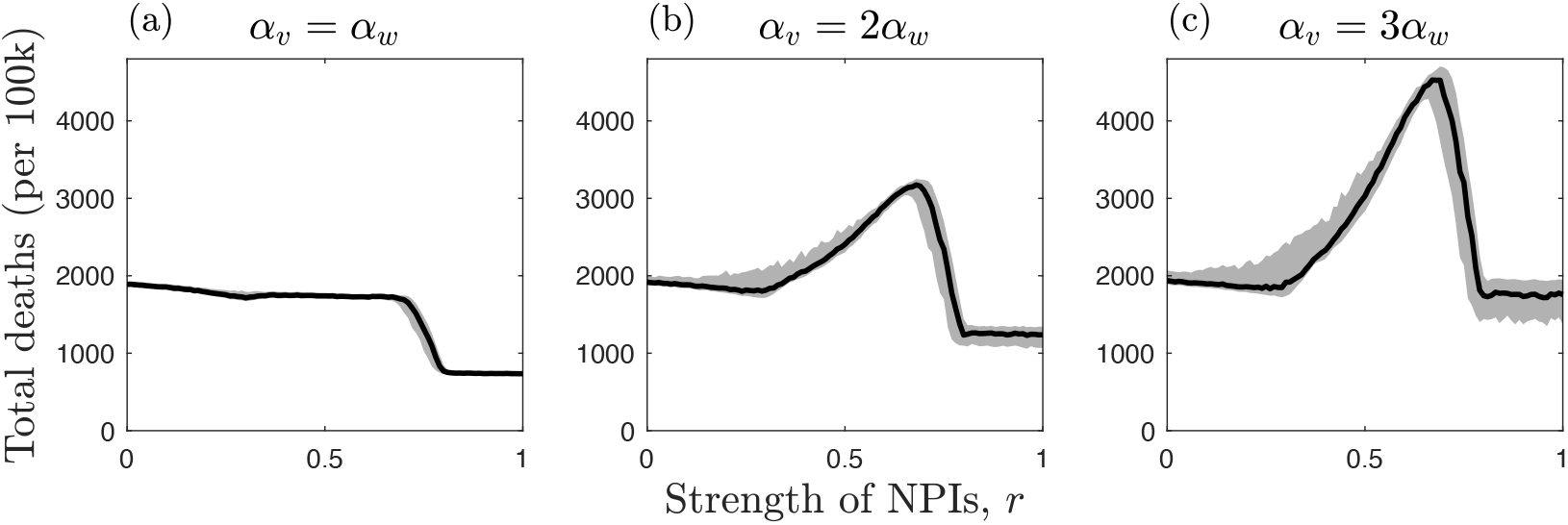
Effects of NPIs (*r*) and the relative virulence (*α*_*ν*_/*α*_*w*_) of a variant that is twice as transmissible (*β*_*ν*_/*β*_*w*_ = 2) on total deaths (per 100k) when there is full cross immunity (*c* = 1) and NPIs have trigger and relaxation thresholds (*∈*_*on*_ = 0.01 and *∈*_*off*,,_ = 0.002). Plots show the median number of deaths (black) along with the upper and lower quartiles (grey). Other parameters as described in the main text with 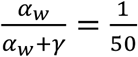.

## Discussion

The emergence of novel variants depends on the interaction between mutation supply and the strength of selection, both of which are influenced by NPIs. Here, we have shown how NPIs and the stage of an epidemic at which they are triggered affect the emergence of novel variants with a range of life-history characteristics. Although stronger, consistent implementation of NPIs generally reduces the likelihood that a novel variant will emerge, more transmissible variants that exhibit a high degree of cross immunity with the wildtype may be most likely to emerge when NPIs are enacted early but are of insufficient strength to drive the wildtype extinct quickly (Fig. 2k, Fig. 3b). This echoes a theoretical result for vaccination, which suggests that imperfect vaccination may provide the optimal conditions for vaccine-escape variants to emerge (van Boven *et al*., 2005; Saad-Roy *et al*., 2021). This is because imperfect vaccination can allow a significant mutation supply while also exerting selective pressure for vaccine-escape variants.

In the results shown here, when NPIs are weak, a large outbreak of the wildtype can occur. This allows a variant to appear (high mutation supply) but prevents it from spreading widely due to the accumulation of cross immunity in the host population (the variant cannot establish). When NPIs are at an intermediate level, however, there may be sufficient mutation supply to allow the variant to appear but insufficient accumulation of cross immunity from the wildtype. The higher transmissibility of the variant then facilitates its establishment. If the variant is also sufficiently more virulent, then it is possible that this will increase the overall number of deaths (Fig. 4). These patterns are broadly when NPIs have trigger and relaxation thresholds based on disease prevalence (Fig. 5, Fig. 6), but in general the variant is able to emerge over a wider set of conditions compared to when NPIs are introduced at the start of the epidemic and are maintained throughout.

While it is possible that intermediate strength or timely NPIs may occasionally lead to more negative outcomes than weaker or delayed NPIs as described above, we note that this requires the variant to have a rather specific set of characteristics. Since it is challenging to predict the phenotypic characteristics of novel variants, we contend that the optimal strategy to prevent variants of concern arising is almost always to ensure that NPIs are strong, timely and implemented consistently. If this is done, then the mutation supply is constrained over the duration of the epidemic, preventing novel variants from appearing in the first place.

Of course, when deciding to implement strong and timely NPIs, a range of factors must be considered. For example, the wildtype might fade out without invading the host population in the absence of NPIs (Gandon *et al*., 2013; Chabas *et al*., 2018). In that case, it may be unnecessary to introduce costly NPIs (Thompson *et al*., 2018). On the other hand, it may be impossible to contain or eradicate a pathogen, even if strong NPIs are introduced (Thompson *et al*., 2020). In that scenario, potential negative non-disease health outcomes of NPIs should be considered, particularly if NPIs are maintained over long periods.

Comparing the costs and benefits of a range of public health measures is an essential area of research (Newbold *et al*., 2020; Sandmann *et al*., 2021; Smith *et al*., 2022). As we have demonstrated, potential evolutionary consequences of different NPIs could be an important component of such analyses.

Our study is related to previous work by Hartfield and Alizon (2014), who explored the relationship between the appearance and establishment phases of variant emergence when the wildtype is weakly adapted to the host (*R*_0_ ≈1) and the variant is strongly adapted (*R*_0_ ≫1). Crucially, Hartfield and Alizon (2014) found that when the *R*_0_ of the wildtype is intermediate (but close to 1), then a variant can emerge that has a lower *R*_0_, which is similar to our finding that intermediate NPIs can facilitate variant emergence since reducing the spread of the wildtype leaves a larger pool of susceptible hosts for the variant. Conversely, the fact that greater spread of the wildtype or high levels of vaccination can reduce the likelihood of a variant with high cross-immunity emerging is also well-established in the literature. While Hartfield and Alizon’s model can be readily adapted to account for NPIs by multiplying the reproductive numbers by (1 − *r*), their analytic approach considers scenarios in which *R*_0_ ≈1 for the wildtype and there is full cross-immunity with the variant (*c* = 1). We were unable to use a similar analytic approach to Hartfield and Alizon (2014) due to additional complexities in our model. Specifically, the assumption of less-than-full cross-immunity is particularly challenging from an analytic perspective, as one would need to track both the depletion of fully susceptible hosts (*S*_*t*_) and hosts that have partial cross-immunity 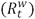.

The present study differed from this previous work in several key aspects. In particular, we explored the probability of variant emergence and the effects of overall mortality when the wildtype has *R*_0_ ≫1 (i.e. the wildtype is likely to cause a large-scale epidemic), when cross-immunity may be partial or absent (*c* < 1), and when NPIs of varying strengths are implemented consistently or with trigger and relaxation thresholds. The differences in our modelling assumptions allowed us to show how interactions between NPIs, cross-immunity, and virulence can mediate not only the emergence of novel variants, but also how under certain circumstances moderate NPIs could theoretically increase overall deaths if a new variant is more virulent than the wildtype.

To explore the potential effects of NPIs on the emergence of pathogen variants, we made several simplifying assumptions in our modelling approach. First, we assumed that there is no differential effect of NPIs on the wildtype and the variant transmission rates. While this is often likely to be true, it is possible that NPIs may affect some strains more than others. For example, if symptomatic people are more likely to be identified and isolated, then selection may favour variants that cause more presymptomatic or asymptomatic infections or that have a longer incubation period. Similarly, different variants may have different generation times (Hart *et al*., 2022), and if individuals are isolated following contact tracing, then variants with shorter generation times may be favoured. Second, our model does not include population heterogeneity or contact structure, both of which affect pathogen transmission and the emergence of variants (Chabas *et al*., 2018). If, for example, some individuals are less likely to adhere to NPIs, then there may be an increased opportunity for new variant appearance. This effect may be particularly pronounced if those individuals belong to specific groups in a population within which transmission may occur. Third, for simplicity we assumed that the wildtype and variant only differed by one mutation at a single genetic locus with potential pleiotropic effects on antigenicity, transmissibility, and virulence. In reality, genetic and phenotypic landscapes are complex, with multiple mutations sometimes required to transition between variants, some of which may be initially neutral or deleterious due to epistasis. For example, several major variants of SARS-CoV-2 exhibit large numbers of mutations, especially in the spike protein (Tegally *et al*., 2021; Volz *et al*., 2021). If a large number of mutations are required to substantially escape host immunity then epistasis may severely constrain pathogen evolution with immunocompromised hosts potentially crucial for antigenic evolution (Smith & Ashby, 2022). Fourth, we assumed that when NPIs were active, they were fixed at a constant level, which may be reasonable if they are in place for a relatively short period of time. However, if one were to consider multi-wave epidemics over a longer period of time (e.g. due to the relaxation of NPIs, seasonality, waning immunity, or pathogen evolution), then it would be more realistic to vary the strength of NPIs as disease prevalence changes rather than just having trigger and relaxation thresholds. It may also be difficult to accurately estimate the current state of the epidemic, especially during the early stages when widespread testing may be unavailable. Finally, we did not model the effects of vaccination programmes on the potential for new variants to emerge, as these have been considered extensively elsewhere (McClean, 1995; Wilson *et al*., 1998; van Boven *et al*., 2005; Day *et al*., 2008; Cobey *et al*., 2021; Gog *et al*., 2021; Saad-Roy *et al*., 2021; Thompson *et al*., 2021).

Despite these simplifications to our model, our results allowed us to demonstrate important principles about the effects of NPIs on the emergence on variants. Our findings are likely to be qualitatively robust with respect to the effects of NPIs, and the framework that we have provided can be readily extended. For instance, some differential effects of NPIs are simply equivalent to altering the transmissibility of the different strains. Population heterogeneity and contact structure both affect pathogen transmission, and so are also likely to affect the initial spread of variants, potentially allowing variants to gain a foothold in a subset of the population. Conversely, a more complex genetic and phenotypic landscape would likely make it more difficult for variants to emerge, for example due to epistasis (Smith & Ashby, 2022). One could crudely model this by reducing the mutation supply in our model to mimic the lower rate of accumulating multiple mutations, which would quantitatively, but not qualitatively, change our results.

In conclusion, NPIs have significant impacts on the emergence of novel variants by mediating both the mutation supply and the strength of selection. Although stronger NPIs generally reduce the probability that a new variant of concern will emerge, there are certain circumstances – namely, when cross immunity is high and the variant is more transmissible – where NPIs of intermediate strength lead to an increased probability of variant emergence, potentially leading to a higher level of mortality. However, this requires a very particular set of circumstances and one cannot predict where a variant will emerge in phenotype space (i.e. its transmissibility, virulence, and level of cross immunity). The optimal strategy to prevent variants emerging is therefore to ensure that NPIs are sufficiently strong to drive the wildtype extinct, thereby cutting off the mutation supply.

## Data Availability

Source code for the simulations is available in the online Supplementary Materials and at https://github.com/ecoevogroup/Ashby_Thompson_2021.

## Author Contributions

BA conceived the study and conducted the investigation. BA and RNT discussed the original ideas and results. CAS carried out additional analyses. BA and RNT wrote the manuscript.

## Data Accessibility

Source code for the simulations is available in the online *Supplementary Materials* and at https://github.com/ecoevogroup/Ashby_Thompson_2021.

## Acknowledgements

BA and CAS are supported by the Natural Environment Research Council (grant numbers NE/N014979/1 (BA) and NE/V003909/1 (BA and CAS)).

