## Supplementary material for "Non-pharmaceutical interventions and the emergence of pathogen variants": Fig. S1

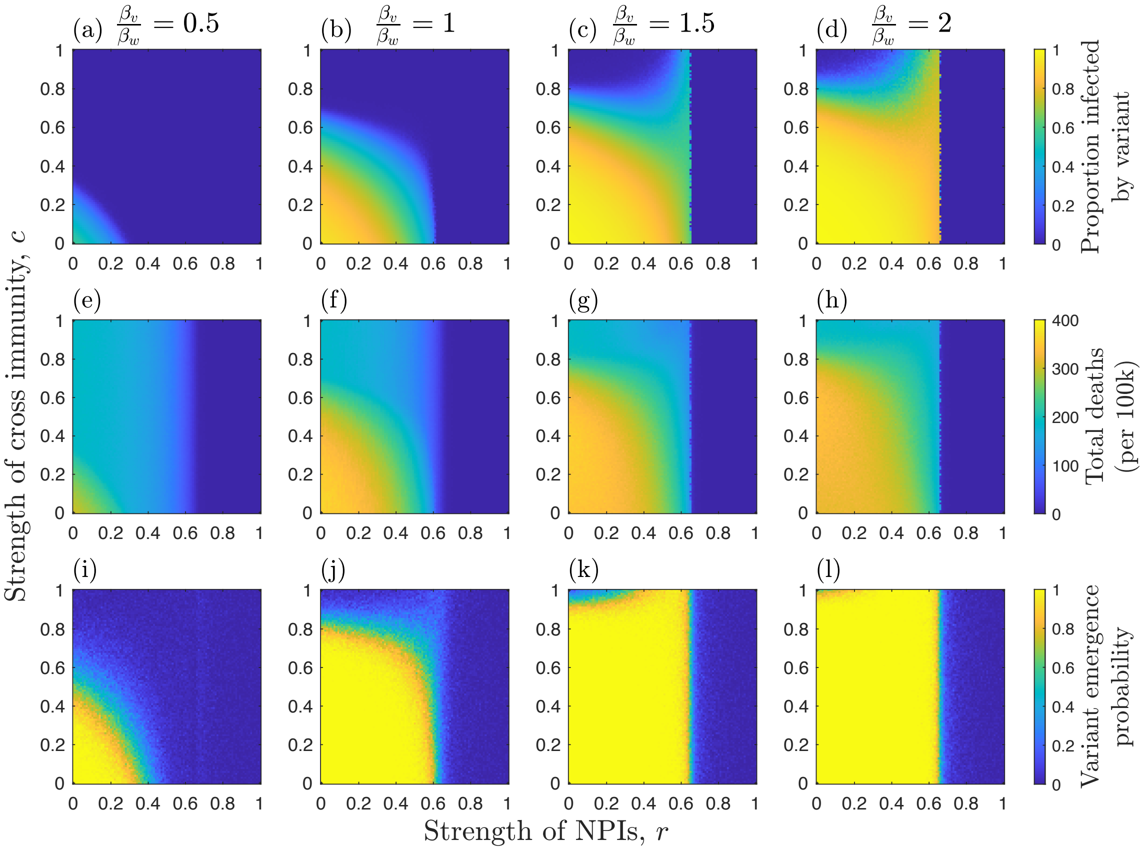


Figure S1 – Effects of the strength of NPIs ($r$), strength of cross immunity ($c$), and relative transmissibility of the variant ($\beta_v/\beta_w$; as indicated at the top of each column) on: (a)-(d) median proportion of hosts infected by the variant; (e)-(h) median total deaths (per 100k) for both strains; and (i)-(l) the probability of the variant emerging (reaching a frequency of at least 0.1). Each measure is calculated over the full duration of each simulation. NPIs are triggered at the start of each simulation and remain in place throughout. Other parameters as described in the main text, with: $\frac{\alpha_w}{\alpha_w+\gamma} = \frac{1}{500}$ and $\alpha_v = \alpha_w$. Results shown for 100 simulations.

**Supplementary material: Non-pharmaceutical interventions and the emergence of pathogen variants**

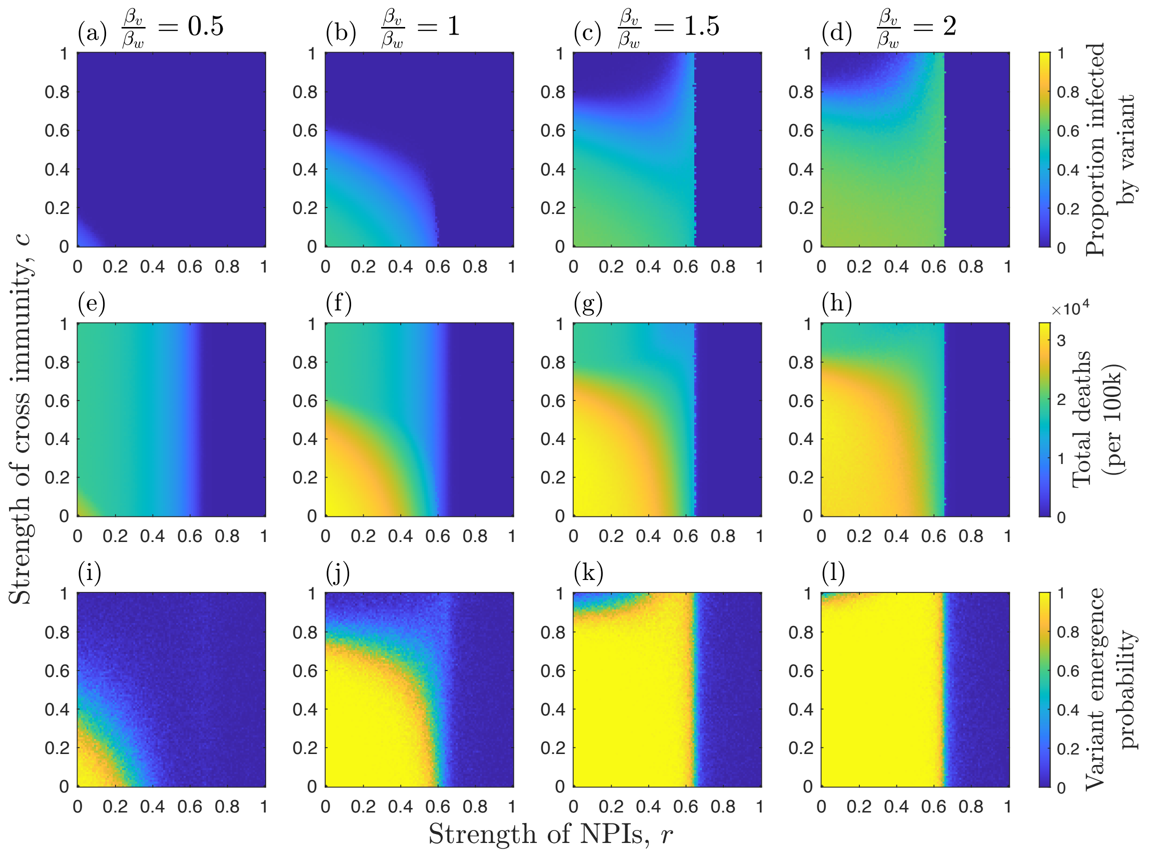


Figure S2 – Effects of the strength of NPIs ($r$), strength of cross immunity ($c$), and relative transmissibility of the variant ($\beta_v/\beta_w$; as indicated at the top of each column) on: (a)-(d) median proportion of hosts infected by the variant; (e)-(h) median total deaths (per 100k) for both strains; and (i)-(l) the probability of the variant emerging (reaching a frequency of at least 0.1). Each measure is calculated over the full duration of each simulation. NPIs are triggered at the start of each simulation and remain in place throughout. Other parameters as described in the main text, with: $\frac{\alpha_w}{\alpha_w+\gamma} = \frac{1}{5}$ and $\alpha_v = \alpha_w$. Results shown for 100 simulations.

**Supplementary material: Non-pharmaceutical interventions and the emergence of pathogen variants**

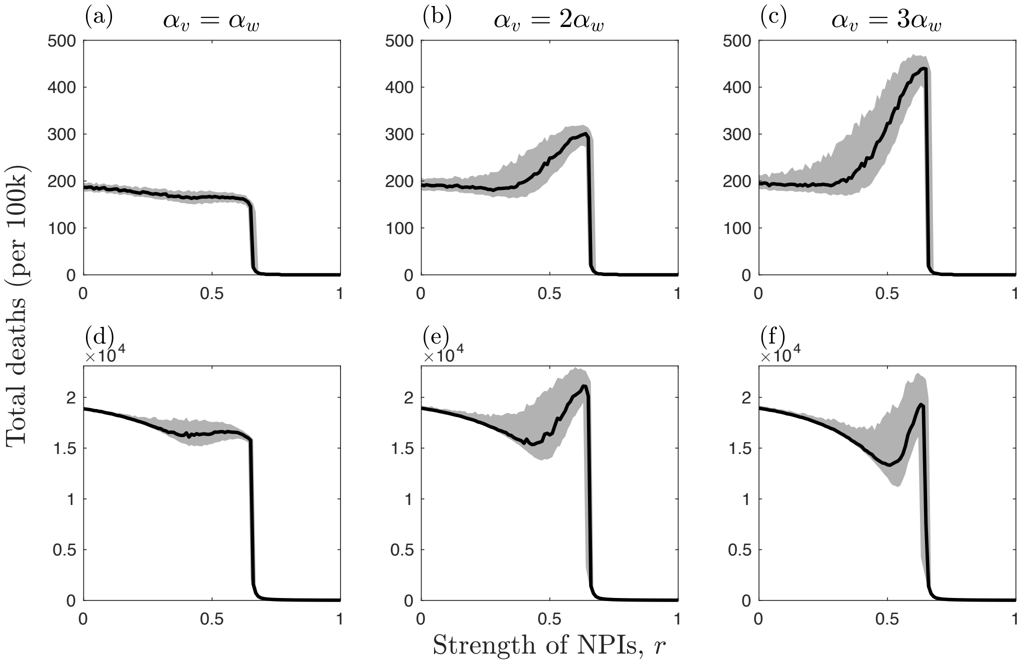


Figure S3 – Effects of NPIs $(r)$ and the relative mortality $(\alpha_v/\alpha_w)$ of a twice as transmissible variant $(\beta_v/\beta_w = 2)$ on total deaths (per 100k) when there is full cross immunity $(c = 1)$ and NPIs are introduced at the start of the simulation and remain in place throughout. Plots show the median number of deaths (black) along with the upper and lower quartiles (grey). Other parameters as described in the main text with: (a)-(c) $\frac{\alpha_w}{\alpha_w+\gamma} = \frac{1}{500}$ and (d)-(f) $\frac{\alpha_w}{\alpha_w+\gamma} = \frac{1}{5}$.

**Supplementary material: Non-pharmaceutical interventions and the emergence of pathogen variants**

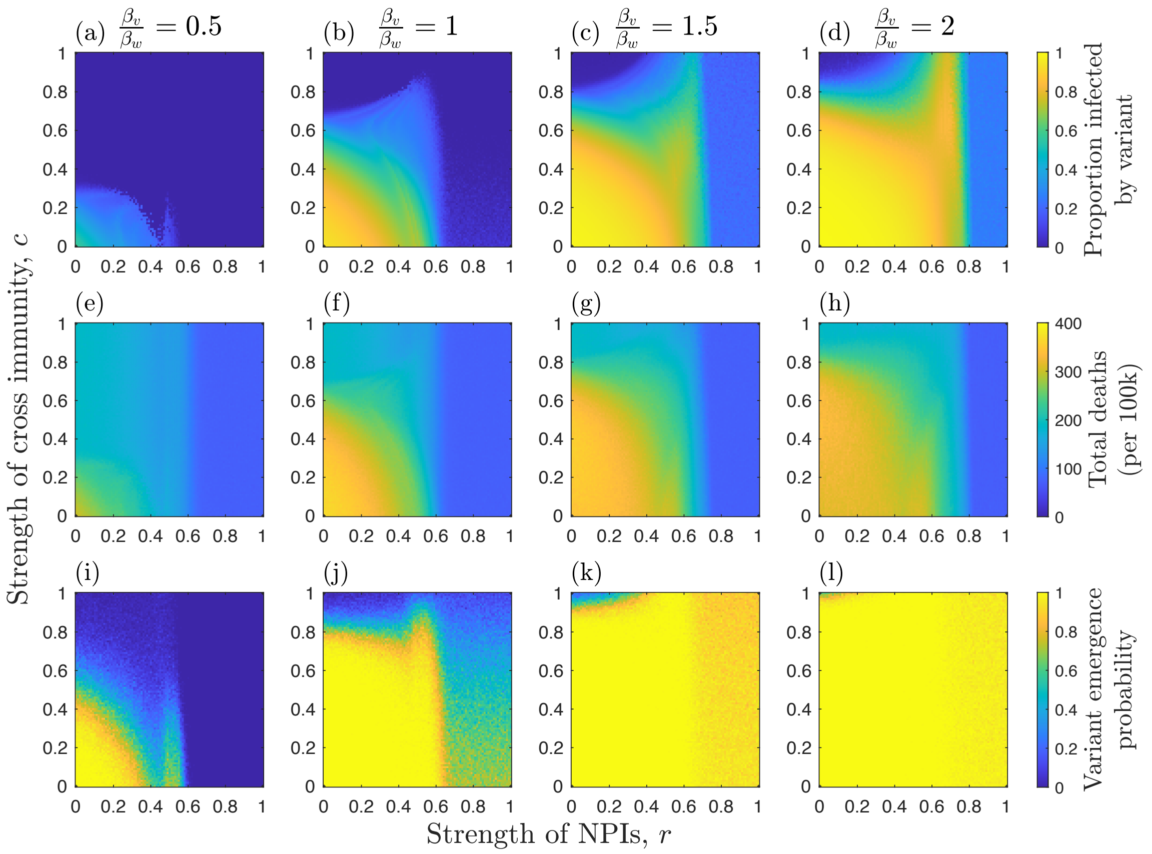


Figure S4 – Effects of the strength of NPIs ($r$) when there are NPI trigger thresholds, strength of cross immunity ($c$), and relative transmissibility of the variant ($\beta_v/\beta_w$; as indicated at the top of each column) on: (a)-(d) median proportion of hosts infected by the variant; (e)-(h) median total deaths (per 100k) for both strains; and (i)-(l) the probability of the variant emerging (reaching a frequency of at least 0.1). Each measure is calculated over the full duration of each simulation. NPIs are triggered at the start of each simulation and remain in place throughout. Other parameters as described in the main text, with: $\frac{\alpha_w}{\alpha_w+\gamma} = \frac{1}{500}$ and $\alpha_v = \alpha_w$. Results shown for 100 simulations.

### Supplementary material: Non-pharmaceutical interventions and the emergence of pathogen variants

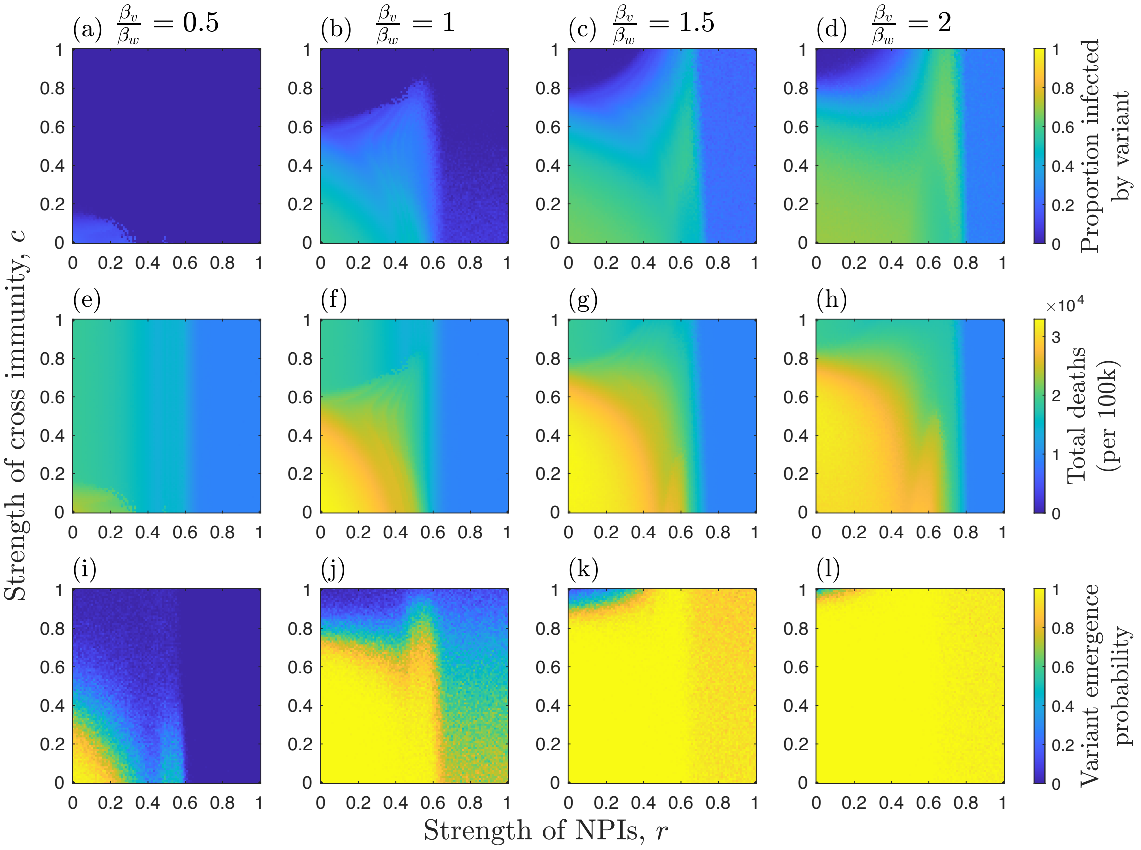


Figure S5 – Effects of the strength of NPIs $(r)$ when there are NPI trigger thresholds, strength of cross immunity $(c)$, and relative transmissibility of the variant ($\beta_v/\beta_w$; as indicated at the top of each column) on: (a)-(d) median proportion of hosts infected by the variant; (e)-(h) median total deaths (per 100k) for both strains; and (i)-(l) the probability of the variant emerging (reaching a frequency of at least 0.1). Each measure is calculated over the full duration of each simulation. The NPI trigger thresholds are $\epsilon_{on} = 0.01$ and $\epsilon_{off} = 0.002$ (i.e. NPIs are triggered when 1% of the host population is infected by either strain, and are removed when only 0.2% are infected). Other parameters as described in the main text, with: $\frac{\alpha_w}{\alpha_w+\gamma} = \frac{1}{5}$ and $\alpha_v = \alpha_w$. Results shown for 100 simulations.

**Supplementary material: Non-pharmaceutical interventions and the emergence of pathogen variants**

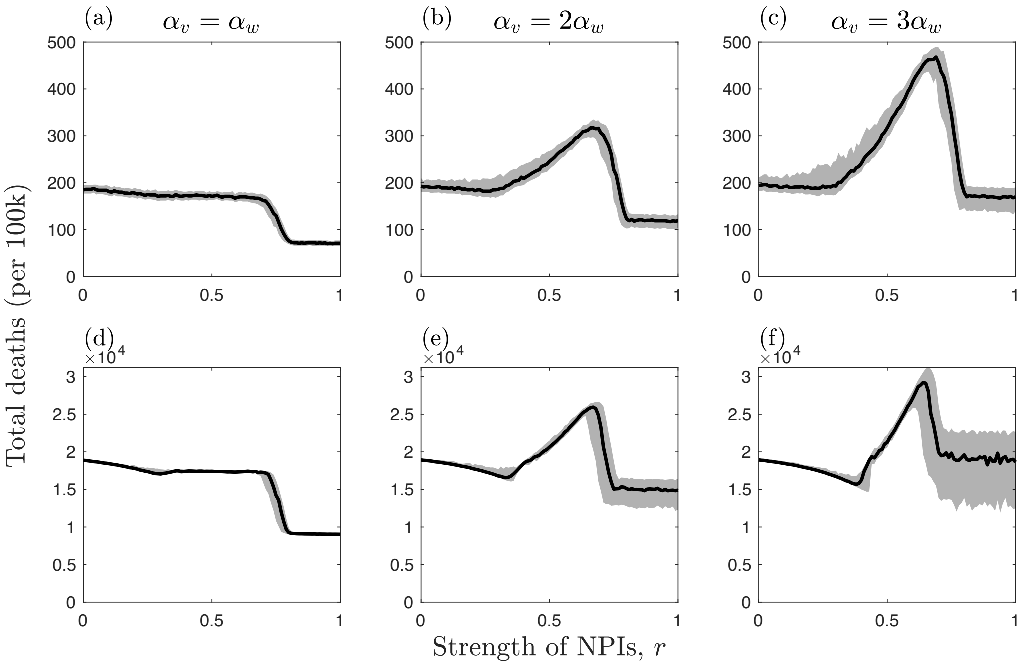


Figure S6 – Effects of NPIs $(r)$ and the relative mortality $(\alpha_v/\alpha_w)$ of a twice as transmissible variant $(\beta_v/\beta_w = 2)$ on total deaths (per 100k) when there is full cross immunity $(c = 1)$ and NPIs have trigger thresholds $(\epsilon_{on} = 0.01, \epsilon_{off} = 0.002)$. Plots show the median number of deaths (black) along with the upper and lower quartiles (grey). Other parameters as described in the main text with: (a)-(c) $\frac{\alpha_w}{\alpha_w+\gamma} = \frac{1}{500}$ and (d)-(f) $\frac{\alpha_w}{\alpha_w+\gamma} = \frac{1}{5}$.
